# Analysis of exome sequencing data implicates rare coding variants in *STAG1* and *ZNF136* in schizophrenia

**DOI:** 10.1101/2024.10.14.24314843

**Authors:** Sophie L Chick, Peter Holmans, Detelina Grozeva, Rebecca Sims, Julie Williams, Michael J Owen, Michael C O’Donovan, James T R Walters, Elliott Rees

## Abstract

Rare coding variants across many genes contribute to schizophrenia liability, but they have only been implicated in 12 genes at exome-wide levels of significance. To increase power for gene discovery, we analysed exome-sequencing data for rare coding variants in a new sample of 4,650 schizophrenia cases and 5,719 controls, and combined these with published sequencing data for a total of 28,898 cases, 103,041 controls and 3,444 proband-parent trios. Novel associations were identified for *STAG1* and *ZNF136* at exome-wide significance and for six additional genes at a false discovery rate of 5%. Among these genes, *SLC6A1* and *KLC1* are associated with damaging missense variants alone. Four of the eight novel genes are also enriched for rare coding variants in other developmental and psychiatric disorders. Moreover, *STAG1* and *KLC1* have fine-mapped common variant signals in schizophrenia. These findings provide novel insights into the neurobiology of schizophrenia, including an aetiological role for disrupted chromatin organisation.

## Introduction

Schizophrenia is a severe and heterogeneous psychiatric syndrome, characterised by behavioural and cognitive symptoms which may be lifelong and are often inadequately resolved by antipsychotic medications^1,2^. Molecular genetic studies have established that the genetic architecture of schizophrenia is highly polygenic, with thousands of alleles of different frequencies from across the genome contributing to liability^3^. Common variants (minor allele frequency (MAF) > 0.01) measured on single-nucleotide polymorphism (SNP) genotyping arrays currently explain 24% of the variance in schizophrenia liability in European populations, and the largest genome-wide association study (GWAS) to date has identified 287 common variant loci at genome-wide significance (mean odds ratio (OR) 1.06; range 1.04–1.23)^4^. Statistical and functional fine-mapping approaches deployed in that report prioritised 106 credible causal protein-coding genes within the associated loci^4^, although few of these genes can be considered definitively implicated.

Current estimates suggest that around 5% of variance in schizophrenia liability is explained by rare (MAF < 0.01) copy number variants (CNVs) and ultra-rare (MAF < 2 x 10^−5^) coding variants^3,5^. To date, 13 rare CNVs have been identified as schizophrenia risk factors, with odds ratios ranging between 1.8 and 81.2^6,7^. All but one of these overlap multiple genes, making it difficult to implicate causal genes. The exception involves non-recurrent, intragenic deletions of *NRXN1*, which encodes a pre-synaptic cell adhesion protein.

Rare coding variants (RCVs) contributing to schizophrenia are concentrated among a set of around 3,000 genes known to be under selective constraint in humans against stop-gain, essential splice site, and frameshift mutations, collectively termed protein-truncating variants (PTVs)^8–11^. The Schizophrenia Exome-Sequencing Meta-Analysis (SCHEMA) consortium analysed data from 24,248 cases, 97,322 controls and 3,402 trios, and identified a set of 10 exome-wide significant genes, with odds ratios ranging between 3 and 50, and a further 22 genes at a false discovery rate (FDR) of < 5%^12^. A subsequent study by the Psychiatric Genomics Consortium (PGC) sequenced the coding regions of 161 genes in 11,580 cases and 10,555 controls, and meta-analysed the gene-level P-values for PTVs with those published by SCHEMA to implicate two additional genes, *AKAP11* and *SRRM2*, at exome-wide significance^13^. Among the 12 genes currently implicated in schizophrenia at exome-wide significance, 5 are also enriched for RCVs in other developmental disorders (DDs), including intellectual disability^12,13^. Moreover, two genes, *GRIN2A* and *SP4*, were prioritised as credible causal candidates in the most recent large schizophrenia GWAS^4^.

The functional roles of genes enriched for RCVs in schizophrenia have informed our understanding of some of the disease mechanisms that may contribute to this disorder. While the genes currently implicated at exome-wide significance have diverse biological functions, several have roles related to glutamatergic synaptic signalling and transcriptional regulation^12,13^. Identifying additional genes enriched for RCVs in schizophrenia will provide further opportunities to understand the complex neurobiology underlying this disorder.

Here, we report whole-exome sequencing data in a new sample of 4,650 schizophrenia cases and 5,719 controls, and combine these with published exome-wide sequencing data for a total of 28,898 cases, 103,041 controls and 3,444 proband-parent trios. In the largest exome-sequencing meta-analysis of schizophrenia to date, we report novel exome-wide significant associations for *STAG1* and *ZNF136* and novel associations at FDR < 5% for six additional genes.

## Results

### New exome sequencing sample

We generated and analysed exome-sequencing data in 4,650 schizophrenia cases and 5,719 controls passing quality control, none of which have contributed to previous exome-sequencing studies of schizophrenia (Supplementary Methods). In this new sample, singleton (minor allele count (MAC) = 1 and absent in gnomAD controls^10^) PTVs, as well as singleton missense variants with ‘missense badness, Polyphen-2 and constraint’ (MPC) scores > 3, in constrained genes were enriched in cases compared with controls (Figure 1, Supplementary Table 4). The effect sizes for these variants in our new sample were consistent with those in the published SCHEMA study (Figure 1)^12^. The new cases were not significantly enriched compared with controls for singleton missense variants with MPC scores between 2 and 3 in constrained genes, but the effect size for these variants was again consistent with the SCHEMA study (Figure 1).

**Figure 1.**
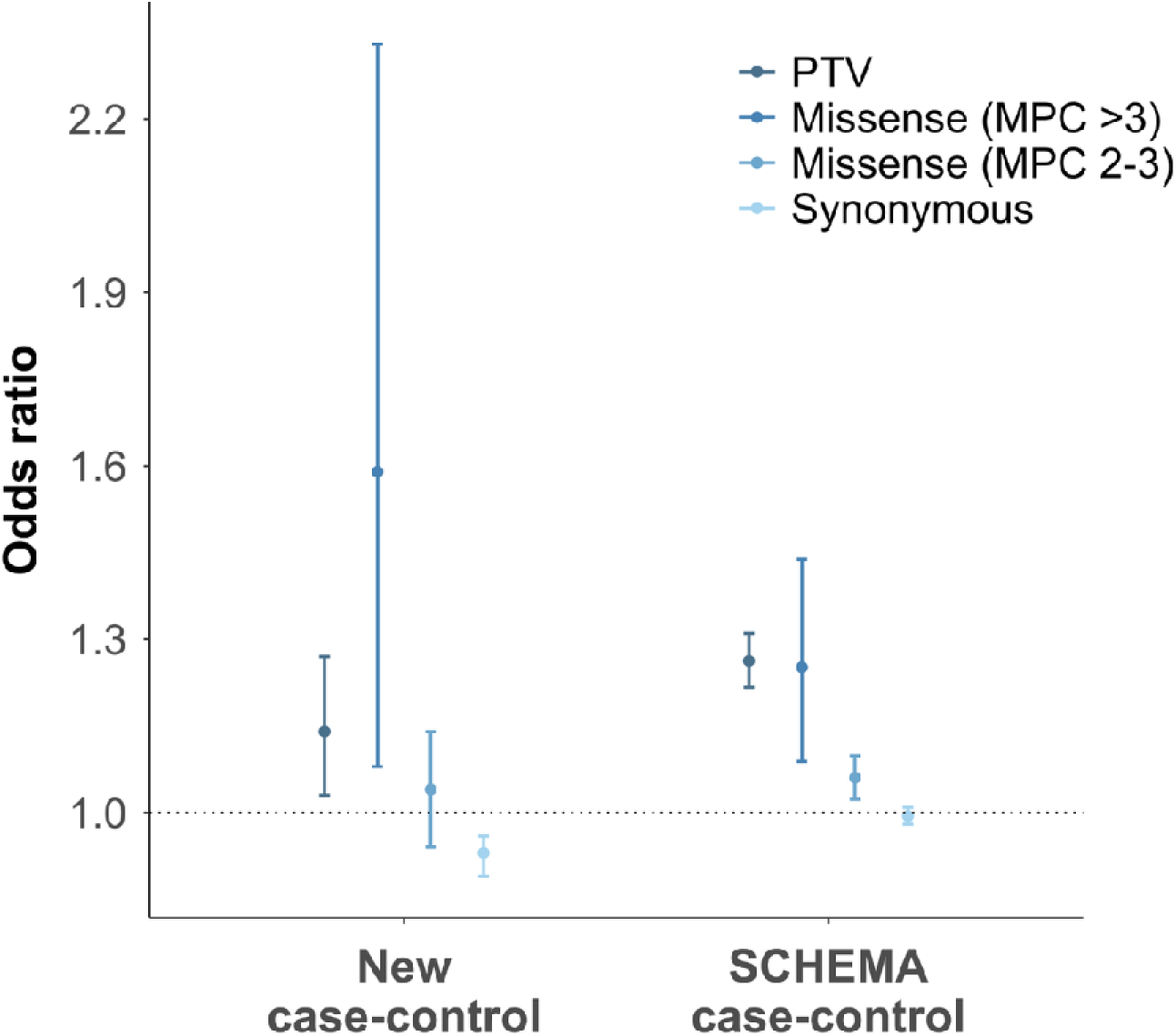
Case-control gene-set analysis of singleton coding variants in constrained genes. Constrained genes are defined as those with pLi scores ≥ 0.9 in gnomAD (n = 3,051)^10^. Odds ratios (OR) in the new case-control sample were derived from Firth’s logistic regression models (Supplementary Methods). Published ORs from the SCHEMA sample were taken from Singh et al 2022. Error bars indicate 95% confidence intervals. PTV = protein-truncating variants; MPC = ‘missense badness, Polyphen-2 and constraint’ score.

The rate of singleton synonymous variants in constrained genes was significantly higher in our new controls than in cases (Figure 1, Supplementary Table 4). This finding is most likely driven by greater sequencing depth in our controls (mean genotype depth = 34.2x) compared with cases (mean genotype depth = 26.0x). Since sequencing depth is positively correlated with the rate of singleton coding variants (Supplementary Material Section 2), any potential bias arising from this will be towards conservative estimates of schizophrenia rare variant enrichment rather than false positive discovery.

### Discovery of novel schizophrenia genes

Joint analysis of RCVs in our new case-control sample with published RCVs from the SCHEMA cases and controls resulted in a total of 28,898 cases and 103,041 controls. We evaluated genes for RCV enrichment using two-sided Cochran-Mantel-Haenszel (CMH) tests, with separate contingency tables for samples grouped by genetically inferred ancestry and exome capture platform (Methods and Supplementary Methods). This analysis focused on variants with a MAC ≤ 5 in all cases and controls (SCHEMA + new sample) and ≤ 5 in 60,146 gnomAD controls^10^, which is similar to the MAC threshold used in the SCHEMA study^12^. Single gene enrichment tests were performed for four variant classes: PTVs; PTVs + missense MPC > 3; PTVs + missense MPC > 2; missense MPC > 2. Following the approach used in the SCHEMA study^12^, genes achieving a case-control CMH P-value < 0.01 were meta-analysed with gene-based *de novo* coding variant P-values derived from 3,444 published schizophrenia trios^11^, using Fisher’s combined probability tests. In total, 30,674 single gene tests were performed, corresponding to an exome-wide significance threshold of P = 1.63 x 10^-6^ after Bonferroni correction. See Methods for a full description of our gene discovery approach.

We identified two novel risk genes at exome-wide significance (Table 1): *STAG1* was associated with rare PTVs and missense (MPC > 2) variants (P = 3.7 x 10^-8^), and *ZNF136* was associated with rare PTVs (P = 6.2 x 10^-7^). We additionally implicated six novel risk genes at FDR < 5% (Table 1). Among these, *SLC6A1* and *KLC1* are associated with schizophrenia based on evidence from missense variants (MPC > 2) alone. In an earlier study^11^, we reported that cases were enriched for *de novo* missense variants (MPC > 2) in *SLC6A1* (3 observed, 0.069 expected, P = 5.2 × 10^−5^). The present study, with inclusion of case-control data, now provides strong support for association between missense variants in *SLC6A1* and schizophrenia (Table 1).

**Table 1.** Novel exome-wide significant and false discovery rate < 5% genes. Odds ratios (ORs) and P-values correspond to the variant class shown. Bold text indicates P-values exceeding Bonferroni significance (P < 1.63 x 10^-6^). P-values are two sided. Q-values show adjusted P-values using the false discovery rate approach. CI = confidence interval; PTV = protein-truncating variant; MPC = ‘missense badness, Polyphen-2 and constraint’ scores for missense variants.

| Gene symbol | Variant class | Combined case-control analysis |  | Trios | Case-control-*de novo* variant meta-analysis |  |
| --- | --- | --- | --- | --- | --- | --- |
|  |  | OR (95% CI) | P-value | *De novo* variants | P-value | Q-value |
| **STAG1** | PTV + missense MPC>2 | 5.6 (2.9-10.6) | **$3.7 \times 10^{-8}$** | 0 | **$3.7 \times 10^{-8}$** | $1.4 \times 10^{-4}$ |
| **ZNF136** | PTV | 4.4 (2.4-8.1) | **$6.2 \times 10^{-7}$** | 0 | **$6.2 \times 10^{-7}$** | 0.0016 |
| **SLC6A1** | Missense MPC>2 | 2.9 (1.5-5.6) | $2.2 \times 10^{-3}$ | 3 | $1.9 \times 10^{-6}$ | 0.0035 |
| **PCLO** | PTV | 4.5 (2.3-8.6) | $2.7 \times 10^{-6}$ | 0 | $2.7 \times 10^{-6}$ | 0.0043 |
| **ZMYND11** | PTV | 7.8 (2.0-29.5) | $7.9 \times 10^{-5}$ | 1 | $1.6 \times 10^{-5}$ | 0.017 |
| **BSCL2** | PTV | 4.5 (2.1-9.4) | $1.9 \times 10^{-5}$ | 0 | $1.9 \times 10^{-5}$ | 0.019 |
| **KLC1** | Missense MPC>2 | 5.0 (2.1-11.9) | $5.0 \times 10^{-5}$ | 0 | $5.0 \times 10^{-5}$ | 0.042 |
| **CGREF1** | PTV | 3.2 (1.8-5.6) | $5.0 \times 10^{-5}$ | 0 | $5.0 \times 10^{-5}$ | 0.042 |

All variants in the new case-control sample which contribute to our novel schizophrenia genes are presented in Extended Data Table 1. The combined case-control and *de novo* variant meta-analysis results for all genes is available in Extended Data Table 2.

**Table 2.** Published rare coding variant enrichment statistics in the novel schizophrenia genes for additional neuropsychiatric disorders. Bipolar disorder association statistics are taken from^18^, ASD association statistics from^19^, DD/ID statistics from^20^, and epilepsy statistics from^17^. ASD association statistics are log_10_ Bayes Factors; all other association statistics are P-values. Schizophrenia NA values correspond to genes without a potential MPC > 2 variant. Dashes (“-“) correspond to either genes not tested in schizophrenia due to low variant numbers (see Methods), or for genes where enrichment statistics were not reported in the given study.

| Gene | Variant Class | Schizophrenia<br>P-value | Bipolar<br>disorder<br>P-value | ASD<br>Log<sub>10</sub> Bayes<br>factor | DD<br>P-value | Epilepsy<br>P-value |
| --- | --- | --- | --- | --- | --- | --- |
| *STAG1* | PTV | $1.9 \times 10^{-4}$ | 1.0 | 0.33 | $8.7 \times 10^{-4}$ | 0.92 |
| | Missense | $5.3 \times 10^{-5}$ | 0.058 | 0.0 | $4.6 \times 10^{-5}$ | 0.47 |
| *ZNF136* | PTV | $6.2 \times 10^{-7}$ | 0.1 | 0.0 | 0.16 | 0.74 |
|  | Missense | NA | 0.21 | 0.0 | 1.0 | - |
| *SLC6A1* | PTV | - | - | 1.1 | $2.3 \times 10^{-8}$ | 0.16 |
| | Missense | $1.9 \times 10^{-6}$ | **0.014** | **10** | $1.0 \times 10^{-14}$ | $9.5 \times 10^{-5}$ |
| *PCLO* | PTV | $2.7 \times 10^{-6}$ | 0.12 | 0.44 | 0.13 | 0.34 |
|  | Missense | NA | 0.058 | 0.0 | 0.31 | - |
| *ZMYND11* | PTV | $1.6 \times 10^{-5}$ | 0.49 | **1.9** | $2.8 \times 10^{-9}$ | 0.72 |
| | Missense | 0.47 | 0.63 | 0.1 | $5.0 \times 10^{-9}$ | 0.20 |
| *BSCL2* | PTV | $1.9 \times 10^{-5}$ | 0.72 | 0.019 | 0.14 | 0.80 |
|  | Missense | NA | 0.51 | 0.0 | 0.22 | - |
| *KLC1* | PTV | 0.98 | 0.50 | 0.0 | 0.18 | - |
| | Missense | $5.0 \times 10^{-5}$ | 0.62 | 0.056 | 1.0 | 0.086 |
| *CGREF1* | PTV | $5.0 \times 10^{-5}$ | 1.0 | **2.9** | 0.071 | 0.27 |
|  | Missense | NA | 0.24 | 0.0 | - | - |

### Overlap of novel genes with schizophrenia common variant loci

Two of our novel genes, *STAG1* and *KLC1,* were members of the broad fine-mapped set of credible causal genes reported in the largest schizophrenia GWAS to date^4^, and narrowly failed to meet the conservative criteria reported by that group for a prioritised gene. *STAG1* was also reported as a likely RCV associated gene, given it was in the FDR < 5% set of the SCHEMA study^12^. Our study strengthens the evidence implicating *STAG1*, with enrichments for rare PTVs and missense (MPC > 2) variants now attaining exome-wide significance. Like *STAG1*, *KLC1* resides within a schizophrenia GWAS locus containing multiple genes (Figure 2), but in that instance, the 95% credible set of causal variants was dispersed across 23 genes^4^; by implicating missense variants in *KLC1* in schizophrenia at FDR < 5%, our study supports the prioritisation of this gene within its GWAS locus.

**Figure 2.**
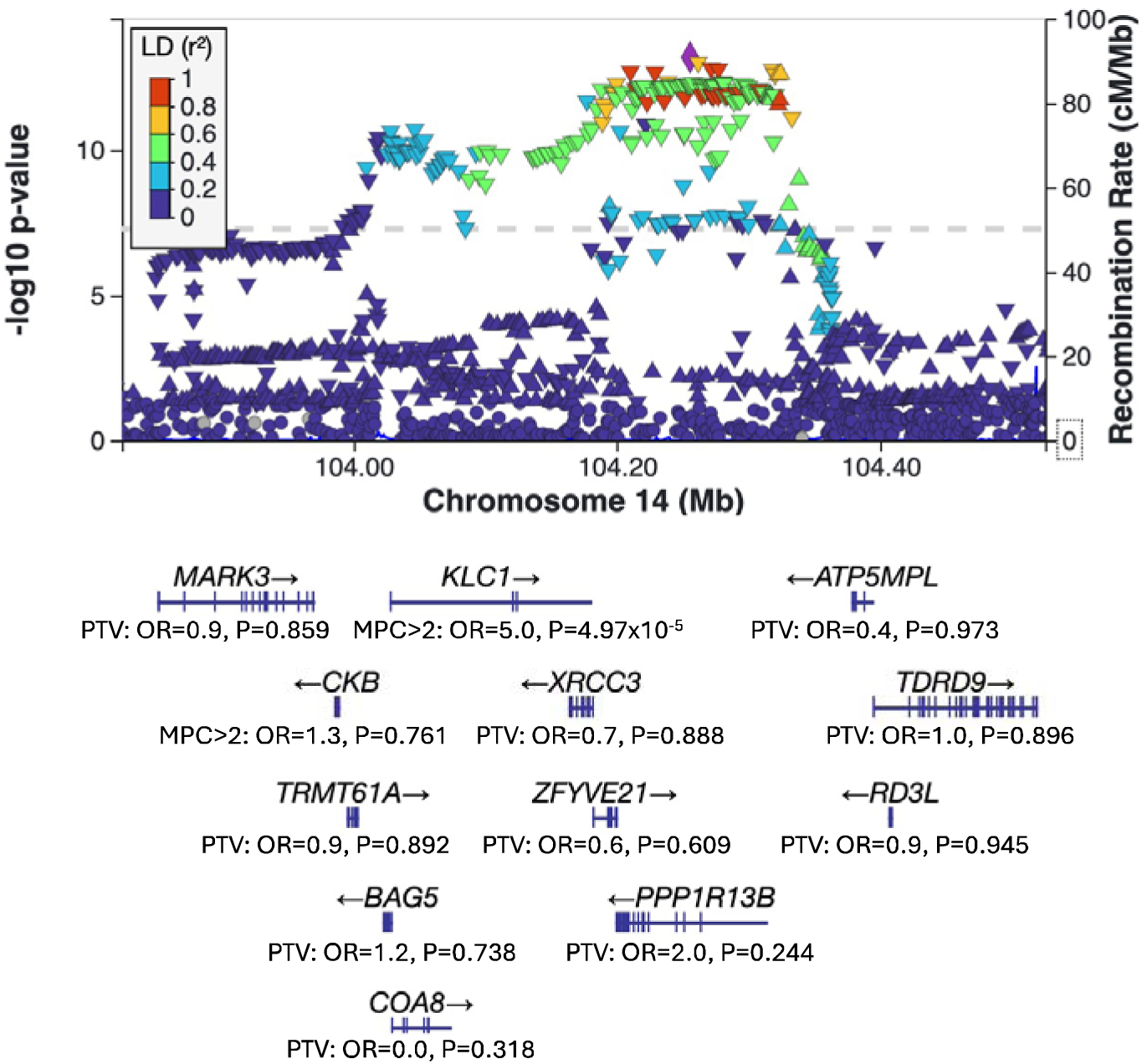
Schizophrenia GWAS locus plot for *KLC1*. Summary statistics for the PGC3 schizophrenia GWAS ^4^ were downloaded from the PGC website (https://pgc.unc.edu/for-researchers/download-results/) and visualised using LocusZoom^14^. For each gene within 200KB of *KLC1*, case-control enrichment P values and odds ratios (OR) from the current study are provided for the class of variant most strongly associated with schizophrenia.

### Pleiotropic effects of novel genes

We examined whether the novel risk genes identified in our study are associated with RCVs in published studies of DD^15^, autism spectrum disorder (ASD)^16^, epilepsy^17^ and bipolar disorder (BD)^18^, and found evidence of pleiotropic effects for 4 of the 8 novel schizophrenia genes (Table 2). In *SLC6A1,* missense variants have broad effects across schizophrenia, ASD, BD, DD and epilepsy, and PTVs are additionally associated with DD (Table 2). In *STAG1*, both PTVs and missense variants are associated with schizophrenia and DD (Table 2). Missense variants in *STAG1* also show weak evidence for association with BD (P = 0.058). In *ZMYND11*, PTVs are associated with schizophrenia, DD and ASD, whereas missense variants are also associated with DD (Table 2). Finally, both schizophrenia and autism show evidence for association with PTVs in *CGREF1* (Table 2).

### Replication of previously implicated genes

We performed a replication analysis in our new sample of genes previously associated with RCVs in schizophrenia. When tested as a set, the 12 previously implicated exome-wide significant genes were enriched for rare PTVs (MAC ≤ 5 in the new sample and ≤ 5 in 60,146 gnomAD controls^10^) in the new cases compared with controls (OR = 4.93, P = 2.8 x 10^-4^; Supplementary Table 5). The 20 additional previously implicated genes at FDR < 5% were also enriched for rare PTVs in cases (OR = 2.46, P = 1.6 x 10^-2^; Supplementary Table 5). The effect sizes for PTVs in these previously implicated gene sets were significantly greater than the general effects observed for PTVs in constrained genes (Supplementary Table 6). The burden of rare missense variants and synonymous variants in previously implicated gene sets did not differ between the new cases and controls (Supplementary Table 5).

We then performed single-gene enrichment tests in our new sample alone for the 12 genes previously implicated in schizophrenia at exome-wide significance. Eight of the 12 genes had a higher burden of RCVs in our new cases compared to controls (OR > 1), three with a one-sided P-value ≤ 0.05 (Supplementary Table 7).

## Discussion

To discover novel schizophrenia risk genes, we generated exome-sequencing data on a new sample of 4,650 cases and 5,719 controls, and jointly analysed these data with RCVs from published sequencing studies of schizophrenia for a total of 28,898 cases, 103,041 controls and 3,444 trios. This represents, to our knowledge, the largest whole-exome sequencing study of schizophrenia to date.

We report association at exome-wide significance between rare PTVs and damaging missense variants in *STAG1* and schizophrenia. *STAG1* encodes a subunit of cohesin, a protein complex required for sister chromatid cohesion during mitosis and meiosis^21^. Defects in cohesin subunits and interactors are observed in cancer^22^ and in a heterogeneous class of neurodevelopmental disorders termed cohesinopathies, whose pathology is thought to be mediated by a further role of cohesin in 3D genome organisation^23^. Cohesin participates in both chromatin looping^24^ and formation of topologically associating domains (TADs)^25^. Mutations in cohesin components, including *STAG1,* produce disrupted patterns of chromatin contact and gene expression, which have been demonstrated to impact neuronal maturation^26–28^. Previous sequencing studies of schizophrenia have provided additional evidence for dysregulated chromatin in schizophrenia, by showing cases are enriched for RCVs and *de novo* coding variants in sets of genes related to chromatin modification and organisation^12,29^. Moreover, schizophrenia cases carry an excess of rare structural variants disrupting TAD boundaries compared with controls^30^. By implicating *STAG1* at exome-wide significance in schizophrenia, we contribute further evidence suggesting an aetiological role for disrupted chromatin organisation in this disorder.

We also found that rare PTVs in *ZNF136* are enriched in schizophrenia at exome-wide significance. *ZNF136* encodes a zinc-finger protein that contains a Krüppel-associated Box (KRAB) domain, which is thought to act as a transcriptional repressor^31,32^. However, the functional roles of *ZNF136* are not well characterised. Unlike all remaining genes currently shown to be enriched for PTVs in schizophrenia with exome-wide significance, *ZNF136* displays no evidence for selective constraint against PTVs (gnomAD probability of being loss-of-function intolerant = 0)^10^. Small-scale transcriptomic studies suggest that *ZNF136* is downregulated in schizophrenia^33,34^, but the mechanisms by which PTVs in *ZNF136* may increase risk for schizophrenia are unclear.

We identified novel RCV associations at FDR < 5% for 6 further genes (*SLC6A1*, *PCLO*, *ZMYND11*, *BSCL2*, *KLC1* and *CGREF1*). *SLC6A1* and *KLC1* are the first genes to be implicated in schizophrenia at FDR < 5% by missense variants (MPC > 2) alone. Sequencing data included in the published SCHEMA study supports association between damaging missense variants (MPC > 2) in these genes and schizophrenia, but the SCHEMA analysis down-weighted association statistics for missense MPC 2-3 variants relative to PTVs and missense MPC > 3 variants, resulting in lower power to detect associations of this class^12^. *SLC6A1* encodes a gamma-aminobutyric acid (GABA) transporter (GAT-1), which is highly expressed in GABAergic neurons and mediates uptake of GABA from the synaptic cleft of inhibitory synapses. Recently published *in vitro* GABA uptake assay data has provided evidence that some of the published schizophrenia *SLC6A1* missense variants, as well as two of the three missense variants reported in our new cases, confer loss-of-function effects on GAT-1 protein leading to reduced GABA uptake^35^. Thus, our study supports the hypothesis of haploinsufficiency being the disease mechanism underlying risk for schizophrenia from missense variants in *SLC6A1*.

The other missense variant enriched gene, *KLC1,* encodes a light chain subunit of kinesin, a tetrameric protein complex responsible for intracellular transport along the cytoskeleton. Knockdown of *KLC1* in a human cell line has been found to impair neuronal differentiation^36^, however, the functional impact of *KLC1* missense variants observed in schizophrenia risk are unknown. A more detailed overview of known biological functions for the novel FDR < 5% implicated genes is provided in Supplementary Material Section 3.

Previous studies have identified four genes which show both fine-mapped common variant signals in schizophrenia GWAS and an excess of RCVs in cases at either exome-wide significance (*GRIN2A* and *SP4*) or FDR < 5% (*STAG1* and *FAM120A*)^4,12^, providing strong evidence for their role in schizophrenia liability. Our study strengthens the evidence for *STAG1.* It also provides orthogonal support for the prioritisation of *KLC1* as a credible causal gene underlying a complex GWAS signal at this locus. The convergence of common and rare genetic liability in *STAG1* and *KLC1* makes them attractive targets for researchers aiming to develop animal and cellular models of rare high-risk variants, as the common allele signal implies that mechanistic insights gained from these models may have broad relevance across cases.

Genes enriched for RCVs in schizophrenia often exhibit pleiotropic effects for other psychiatric and developmental disorders, particularly DD and ASD^12,37^. Four of the 8 novel genes identified in the current study show enrichment for RCVs in large sequencing studies of DD, ASD and epilepsy, providing orthogonal support for their role in schizophrenia. *SLC6A1* has the broadest pleiotropic effects, wherein missense variants are strongly implicated in ASD, DD and epilepsy, and weakly in BD. PTVs in *SLC6A1* are also associated with DD. Several of the novel genes reported in the current study also demonstrate association with syndromic neurodevelopmental disorders; for example, PTVs, missense variants, and deletions in *STAG1* cause a syndromic cohesinopathy characterised by developmental delay and mild dysmorphic features, sometimes accompanied by autistic traits and epilepsy^38–40^. Homozygous PTVs in *PCLO* are associated with pontocerebellar hypoplasia type III^41^, a cause of global developmental delay and seizures.

Furthermore, loss and gain-of-function mutations in *BSCL2* have been implicated in lipodystrophy and neuropathic conditions respectively^42^. It will be important for future studies to determine whether particular clinical features, including neurodevelopmental phenotypes, are enriched among schizophrenia cases carrying mutations in these pleiotropic genes.

Analysis in the new sample of genes that were previously implicated in schizophrenia at exome-wide significance confirmed this set of genes are enriched for rare PTVs in cases compared with controls. Moreover, while single-gene replication analysis was underpowered, *SETD1A*, *XPO4* and *SRRM2* were enriched for RCVs in the new cases at a nominal significance of P-value < 0.05. However, both our own study and a previous targeted sequencing study^13^ found higher rates of rare PTVs and damaging missense variants in *CACNA1G* in controls than in cases, suggesting further work is required to determine whether *CACNA1G* is a true schizophrenia risk gene.

A strength of our study is the large number of newly exome-sequenced cases, which we combine with published data to increase power for gene discovery. Additionally, the inclusion of a missense only variant test identified two novel genes at FDR < 5% significance. Our study also has limitations. We lack deep and longitudinal phenotype data for most of the new and published cases included in our analysis, and we are therefore unable to determine whether variants in the novel genes reported are associated with particular clinical features. Moreover, we are underpowered to analyse individuals in the new sample from diverse populations. Increasing the diversity of sequenced samples in schizophrenia will both facilitate genomic discovery and ensure more equitable progress in precision psychiatry.

In conclusion, our study implicates *STAG1* and *ZNF136* in schizophrenia with exome-wide significance and 6 additional genes at FDR < 5%. Many of these genes are enriched for RCVs in DD, ASD and epilepsy, which supports their association with schizophrenia given the known genetic overlap between these disorders. We strengthen the evidence for an allelic series of common and rare schizophrenia risk alleles in *STAG1*, and provide novel evidence for the convergence of common and rare risk alleles in *KLC1*. Association of *STAG1* at exome-wide significance implicates disrupted chromatin organisation in schizophrenia, while association of *SLC6A1* at FDR < 5% furthers the evidence for an aetiological role of perturbed GABAergic neuronal signalling.

## Methods

### New case-control exome-sequencing sample

A total of 5,525 blood-derived DNA samples from schizophrenia cases were selected for exome sequencing. The majority of cases (n = 4,482) were from the treatment-resistant schizophrenia CLOZUK cohort^43^. The remaining 1,043 cases came from clinically ascertained cohorts and meet DSMIV^44^ or ICD10^45^ criteria for schizophrenia or schizoaffective disorder. See Supplementary Methods for further details. Our new sample consisted of 7,268 controls before quality control, which were derived from the following collections: Welcome Trust Case-Control Consortium (WTCCC) 2 cohort (n = 1,595)^46^; National Centre for Mental Health (NCMH) cohort (n = 398)^47^; Cardiff Alzheimer’s disease cohort (n = 5,275)^48,49^. Further details about the new control sample are included in the Supplementary Methods.

Case and control recruitment was carried out in line with ethical regulations and with approval from local ethics committees (Supplementary Methods).

### Generation of whole-exome sequencing data

All participants in the new sample were exome-sequenced on the Illumina HiSeq 4000 platform, using the Nextera DNA Exome Capture Kit, Illumina HiSeq 3000/4000 PE Cluster Kit and Illumina HiSeq 3000/4000 SBS Kit. Processing of raw sequence reads was implemented in accordance with Genome Analysis ToolKit (GATK) Best Practice guidelines^50^. The Burrow–Wheeler Aligner (v0.7.15)^51^ was used to align reads to the human reference genome (GRCh37). Variants were joint-called across all samples using the GATK Haplotype Caller V3.4 and filtered using the GATK Variant Quality Score Recalibration (VQSR) tool. Mean genotype depth was 26.0x for cases and 34.2x for controls.

### Sample, variant and genotype quality control

Sample quality control is fully described in Supplementary Methods. Briefly, samples were excluded if they had a call rate < 0.75, a mean genotype depth < 10, or if their sex predicted from the sequencing data did not match their recorded sex. Related individuals were also excluded to ensure that no two samples were third-degree or closer in relationship, prioritising the retention of schizophrenia samples. Principal component analysis was used to assess and control for population structure. Finally, we excluded samples that failed one or more hard sequencing quality filters (described in Supplementary Methods), and also samples overlapping with the SCHEMA study. Following sample quality control, 4,650 cases and 5,719 controls were retained.

### Association analysis

RCVs from the new case-control sample were jointly analysed with published RCVs from the SCHEMA study, resulting in a total of 28,898 cases and 103,041 controls. Autosomal and pseudoautosomal genes were analysed using two-sided Cochran-Mantel-Haenszel (CMH) tests with continuity correction, with separate contingency tables for the 11 SCHEMA strata (Supplementary Table 2) and the new case-control sample. Since sex-stratified variant counts are not available from SCHEMA, association statistics for non-pseudoautosomal genes on the X chromosome were generated using two-sided CMH tests with continuity correction, with separate contingency tables for the 11 SCHEMA strata and 2 additional contingency tables for males and females in the new sample.

We analysed variants with a MAC ≤ 5 in all cases and controls (SCHEMA + new sample). Since some variants in the new sample may not have been included in the SCHEMA study for quality or technical reasons, we also excluded variants in our new sample that had a MAC > 5 in 60,146 gnomAD controls. In our full case-control meta-analysis, per-gene CMH P-values for rare synonymous variants followed a null distribution (Extended Data Figure 1).

Single gene enrichment tests were performed for four variant classes: PTVs (18,318 genes); PTVs + missense MPC > 3 (2,034 genes); PTVs + missense MPC > 2 (4,991 genes); and missense MPC > 2 (4,991 genes). Tests involving missense variants were limited to genes containing at least one possible missense variant with an MPC score above a given threshold (i.e. 2,034 genes have ≥ 1 possible missense variant with an MPC score > 3). To mitigate type I error for genes that only included variants observed in a single stratum with a small number of cases and/or controls, we only tested genes with variants observed in at least two strata, or in the largest European stratum (n gene tests excluded across the four variant classes = 1,724); we note that these excluded tests still contribute to our multiple testing criteria. We also excluded three genes (*TET2*, *DNMT3A,* and *ASXL1*) known to be associated with age-related clonal haematopoiesis (ARCH)^52,53^. In total, 30,334 single gene tests were performed in the case-control analysis.

Following the approach used in the SCHEMA study, we meta-analysed genes achieving a case-control CMH P-value < 0.01 (n genes = 340) with gene-based *de novo* coding variant P-values derived from 3,444 published schizophrenia trios^11^ using Fisher’s combined probability tests. Gene-based *de novo* coding variant enrichment statistics were generated using Poisson rate ratio tests^11,54^. The final test statistic reported for each gene is the minimum P-value across the following tests: **(1)** case-control only (30,334 tests); **(2)** case-control and *de novo* variant (340 tests). This gave a total of 30,674 tests, corresponding to an exome-wide significance threshold of P = 1.63 x 10^-6^. We note that this serves as a conservative threshold, as the tests are not independent given the overlap of variants in each test.

A gene discovery sensitivity analysis that excludes individuals with Alzheimer’s disease from our new control sample is presented in Supplementary Material Section 5.

### Gene set analysis

Gene set enrichment analyses were conducted using Firth’s logistic regression tests. Here, case-control status was regressed on the number of variants carried in a given gene-set, controlling for the first ten principal components derived from common variants in the exome-sequencing data, sex, and the exome-wide burden of singleton synonymous variants. See Supplementary Methods for further details.

## Data availability

The genetic variants contributing to the novel schizophrenia genes reported in the current study are presented in Extended Data Table 1. Aggregated variant counts at the gene level are provided in Extended Data Table 2. All datasets included in the current study are described in the Methods and Supplementary Material. Exome-sequence data from the new cases are deposited into the SCHEMA consortium. Access to WTCCC2 control biological samples is managed and approved by the WTCCC (https://www.wtccc.org.uk/ccc2/). The Accession number for Sequence data from the Alzheimer’s disease European sequencing project, which includes a subset of the Cardiff Alzheimer’s disease cohort, is: dbGaP (phs000572.v7.p4 (stage 1)). SCHEMA case-control variants included in the current study are available for download through the SCHEMA browser (https://schema.broadinstitute.org/).

## Code availability

The software and code used in this study are described in the Methods. The code used to generate our gene set and single gene results are available at https://github.com/sophie-chick/SZ_gene_discovery.

## Acknowledgments

This work was supported by a UKRI Future Leaders Fellowship Grant to E.R (MR/T018712/1), a MRC Program Grant no. MR/P005748/1 to M.J.O, M.C.O’D., J.T.R.W., and P.H., a MRC Program Grant no. MR/Y004094/1 to M.J.O, M.C.O’D., J.T.R.W., P.H and E.R, and a Fieldrose Charitable Trust PhD scholarship to S.L.C, funded by Mental Health Research UK and the Schizophrenia Research Fund. The Cardiff Alzheimer’s disease cohort was supported by an MRC programme grant, funding from Health and Care Research Wales and a charitable donation from the Moondance Foundation. We acknowledge Jun Han, Joanne Morgan, Lesley Bates, Lucinda Hopkins, Rachel Raybould, Nicola Denning, Alun Meggy and Rachel Marshall at Cardiff University, for laboratory sample management and sequencing. We also acknowledge Mark Einon, at Cardiff University, for support with the use and setup of computational infrastructures, and Antonio Pardiñas at Cardiff University for advice on schizophrenia common variant analysis.

## Competing interests

ER, JTRW, MCO and MJO reported receiving grants from Akrivia Health outside the submitted work. JTRW, MJO and MCO reported receiving grants from Takeda Pharmaceutical Company Ltd outside the submitted work. Takeda and Akrivia played no part in the conception, design, implementation, or interpretation of this study.

## Notes

### Author Declarations

The CLOZUK study received UK National Research Ethics Service approval (reference number 10/WSE02/15). The Cardiff COGS study was approved by the South East Wales Research Ethics Committee Panel (reference number: 07/ WSE03/110). The F-series and affected sib studies were approved by the Multicentre Research Ethics Committee in Wales (reference number: MREC 99/9/16). Ethical approval for the NCMH sample obtained from Wales Research Ethics Committee 2 (reference: 16/WA/0323). Ethical approval was obtained from UK NHS Health Research Authority/Health and Care Research Wales and the Wales Research Ethics Committee 3 Cardiff board for the Cardiff Alzheimer's disease cohort (reference numbers 04/9/030 and 17/SS/0139).

