## Supplementary material for "Analysis of exome sequencing data implicates rare coding variants in *STAG1* and *ZNF136* in schizophrenia"

[3.	Summary of novel schizophrenia FDR < 5% genes.	13](#_Toc179790419)

[Supplementary Table 8. Sensitivity analysis of exome-wide significant and FDR < 5% genes.	20](#_Toc179790426)

### 1. Supplementary methods

#### Sample description

**Schizophrenia cases**

4,482 of the new cases were from the CLOZUK2 cohort^1^. Anonymised blood samples from CLOZUK2 were ascertained from patients receiving clozapine and having a clinical diagnosis of treatment-resistant schizophrenia. A validation for using a clinician diagnosis of treatment-resistant schizophrenia against a research diagnostic criteria for schizophrenia can be found in Pardiñas et al 2018^1^, Supplementary Note (2). The new sample also included the following clinically ascertained cohorts: CardiffCOGS (n = 429)^2,3^, F-series (n= 453^4^, and Affected-Sib (n = 161; NB only 1 affected member of each sib-pair was included^5^. All clinically ascertained cases meet DSMIV^6^ or ICD10^7^ criteria for schizophrenia or schizoaffective disorder. Further details on these clinically ascertained samples can be found in the respective published studies cited above.

**Controls**

The new controls were derived from the following cohorts: **1)** 1,595 WTCCC2 controls from the 1958 birth cohort^8,9^; **2)** 398 NCMH controls that were recruited by the National Centre for Mental Health at Cardiff University and screened for the presence of psychiatric disorders^10^. **3)** 5,275 Cardiff Alzheimer’s disease (AD) cohort samples from a recent exome-sequencing project on dementia conducted by Cardiff University. 80% of this sample comprises individuals with Alzheimer’s disease; a subset of these are included in Holstege et al. 2022, which contains further cohort descriptives. We included individuals with AD in our new control sample given no evidence for a significant genetic correlation between AD and schizophrenia^11^.

**Ethics statements**

All research conducted as part of this study was consistent with UK regulatory and ethical guidelines. The CLOZUK study received UK National Research Ethics Service approval (reference number 10/WSE02/15). The Cardiff COGS study was approved by the South East Wales Research Ethics Committee Panel (reference number: 07/ WSE03/110). The F-series and affected sib studies were approved by the Multicentre Research Ethics Committee in Wales (reference number: MREC 99/9/16).

The control samples were recruited as part of independent projects, all of which have equivalent ethical permissions and data sharing procedures in place. Ethical approval for the NCMH sample obtained from Wales Research Ethics Committee 2 (reference: 16/WA/0323). Ethical approval was obtained from UK NHS Health Research Authority/Health and Care Research Wales and the Wales Research Ethics Committee 3 Cardiff board for the Cardiff Alzheimer’s disease cohort (REC numbers 04/9/030 and 17/SS/0139).

#### Sample-level quality control

**Initial sample exclusions and sex checks:** 28 samples were excluded for having a sample call rate < 0.75 or a mean genotype depth < 10. An additional 37 cases and 4 controls were excluded for failing sex checks, where sample sex as imputed using Peddy^12^ did not match their recorded sex. We were unable to perform sex checks on WTCCC2 controls as we did not have access to their recorded sex.

**Relatedness exclusions:** Hail’s PC-Relate method (https://hail.is/docs/0.2/index.html) was used to estimate pairwise kinship coefficients (Φ_ij_) between all pairs of samples, based on linkage disequilibrium (LD) pruned SNPs (max r^2^ < 0.1) with a MAF > 0.01 and a variant call rate > 0.98. The first 2 principal components (PCs) were included to correct for population structure in the kinship calculation. Pairs of individuals where Φ_ij_ **≥** 0.45 were considered duplicates or monozygotic twins, those with Φ_ij_ between 0.1 and 0.4 and identity-by-descent 0 (ibd0) < 0.1 as parent-child relatives, those with Φ_ij_ between 0.1 and 0.4 and ibd0 between 0.1 and 0.4 as siblings, those with Φ_ij_ between 01 and 0.2 and ibd0 between 0.4 and 0.7 as 2^nd^ degree relatives, and those with Φ_ij_ between 0.1 and 0.2 and ibd0 > 0.7 as 3rd degree relatives (Supplementary Figure 1). We excluded related individuals to ensure that no two samples were third-degree or closer in relationship, prioritising the retention of schizophrenia samples. This resulted in the exclusion of 224 cases and 331 controls.

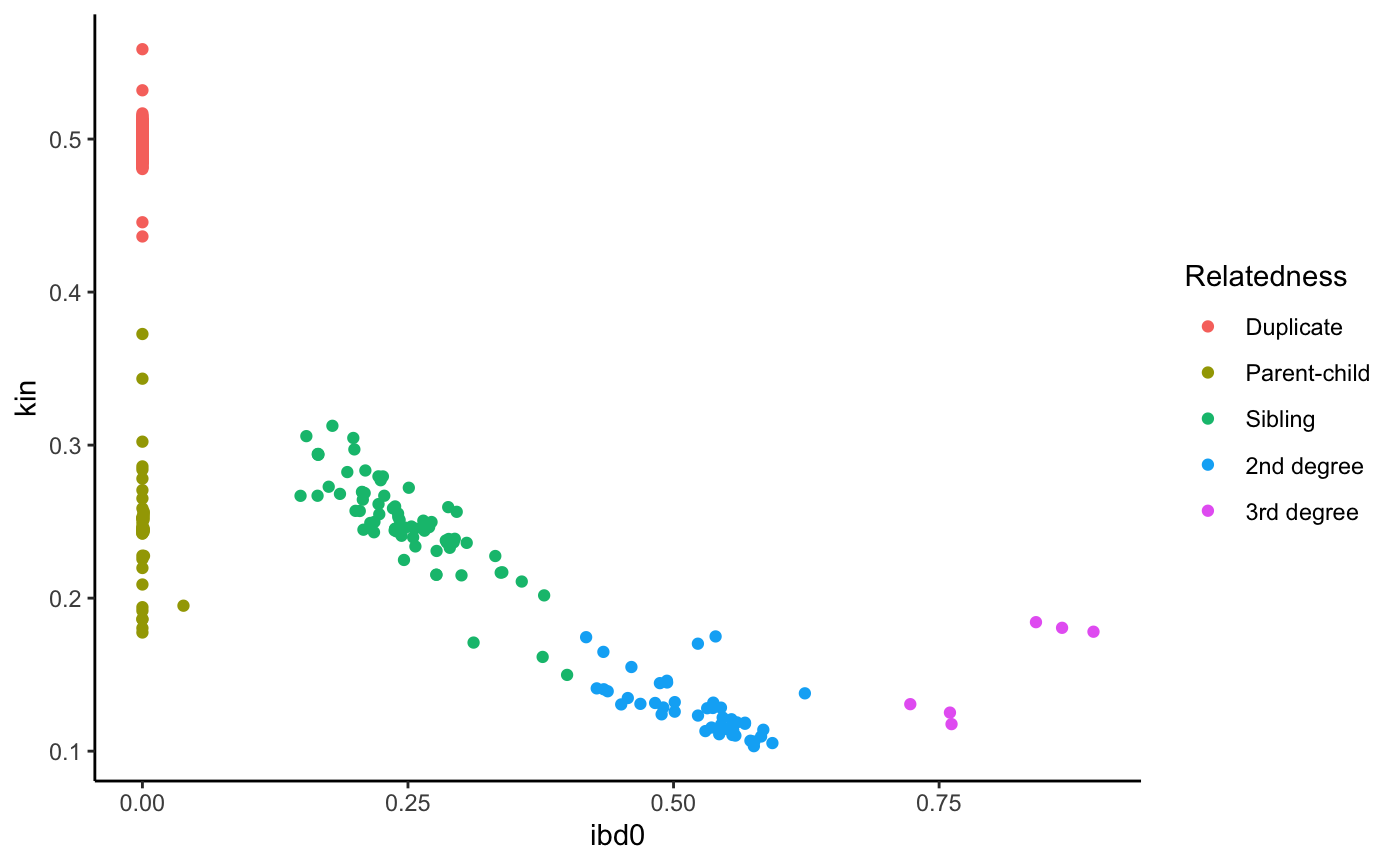

**Supplementary Figure 1.** Sample relatedness in the new case-control dataset. Estimated kinship coefficients (kin) and proportion of identity-by-descent zero (ibd0) alleles for each pair of individuals with a kinship coefficient greater than 0.1. Kin and IBD0 statistics were generated using Hail’s PC-Relate method^13^.

**Controlling for population structure:** Individuals in the new case-control sample were first grouped based on genetic similarity to the superpopulations used in the 1000 genomes (1KG) project. Here, Peddy^12^ was used to perform a principal components (PC) analysis and then train a support vector machine (SVM) on the first 4 PCs derived from samples in the 1KG project, using the 1KG sample superpopulations as training labels (Supplementary Figure 2 A). To control for population structure, we first excluded 140 cases and 36 controls that fell 4 standard deviations or more from the means of PCs 1 and 2 in samples predicted by Peddy to be genetically similar to the 1KG European superpopulation reference (Supplementary Figure 2 A). We then applied Hail’s hwe_normalized_pca function to the remaining samples, which identified two PC-associated clusters (Supplementary Figure 2 B). As shown in Supplementary Figure 2 B, the majority of samples were observed in cluster 1, with cluster 2 containing only 5.5% of cases and 18.1% of controls. The mean number of singleton variants carried by samples within the boundaries of PCA-cluster 1 as shown in Supplementary Figure 2 B (mean n = 88.3; standard error (SE) = 0.23) was less than half of that observed for samples outside of this cluster (mean n = 210.2; SE = 1.55). To ensure our findings are not confounded by population structure, we restricted our analysis to samples within the boundaries of PCA-cluster 1 as shown in Supplementary Figure 2 B. This resulted in the exclusion of an additional 278 cases and 892 controls.

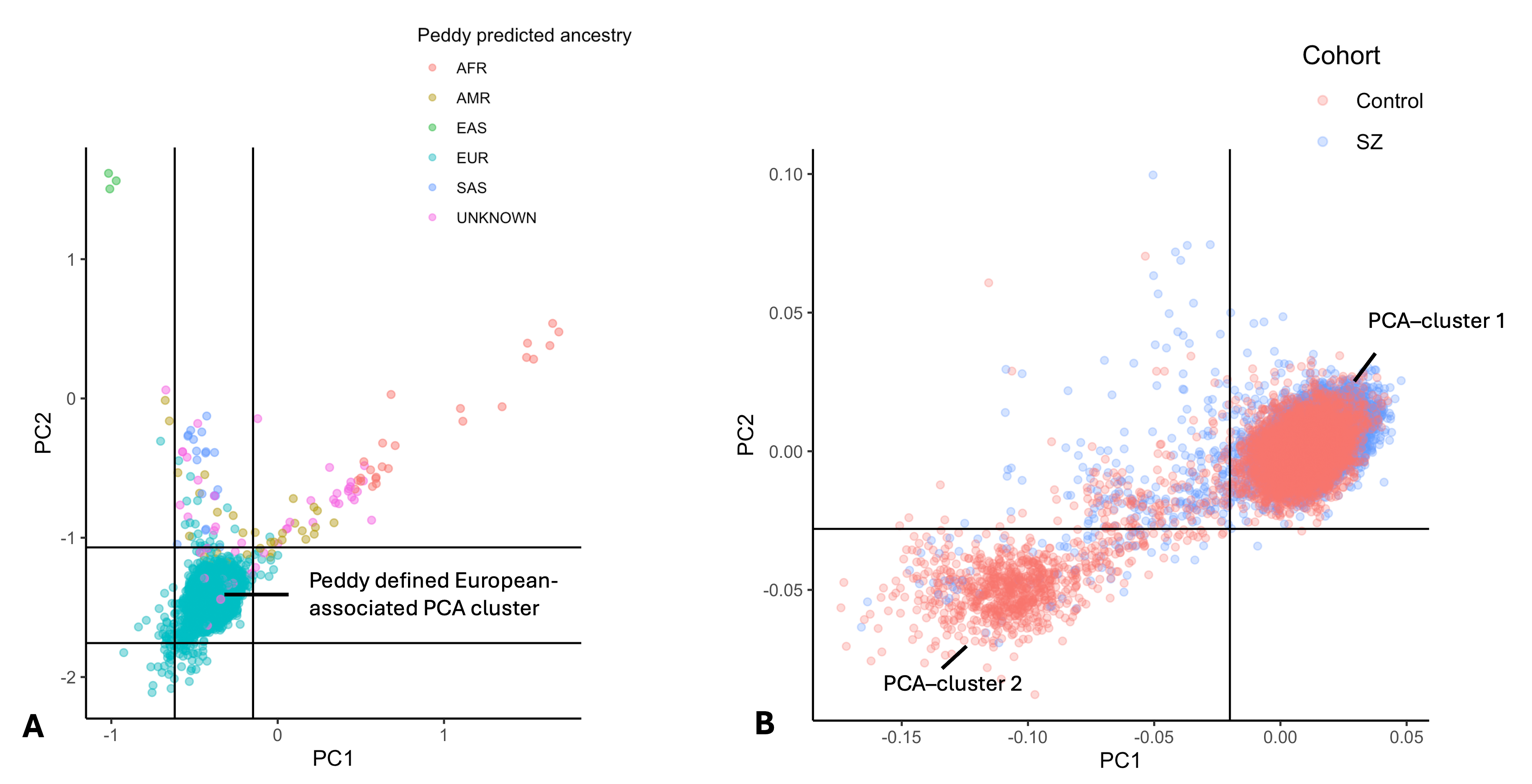

**Supplementary Figure 2. A)** Principal component analysis and sample ancestry as predicted by Peddy. Horizontal and vertical lines indicate 4 standard deviations from the mean of PC1 and PC2 in samples defined by Peddy as belonging to the European-associated PCA cluster. **B)** Principal component analysis for samples in the Peddy-defined European-associated PCA cluster (see Figure 3A). To control for population structure, samples outside of the boundaries of PCA-cluster 1 were excluded.

**Hard filters:** For samples passing all quality control described above, we applied Hail’s sample_qc function to variants passing genotype and variant QC (described below), and excluded samples based on the following filters: call rate < 0.96; number of singletons > 110; TiTV ratio < 2.95 or > 3.15; het/hom ratio < 1.43 or > 1.82 (Supplementary Figure 3). A total of 232 samples were excluded after applying hard filters.

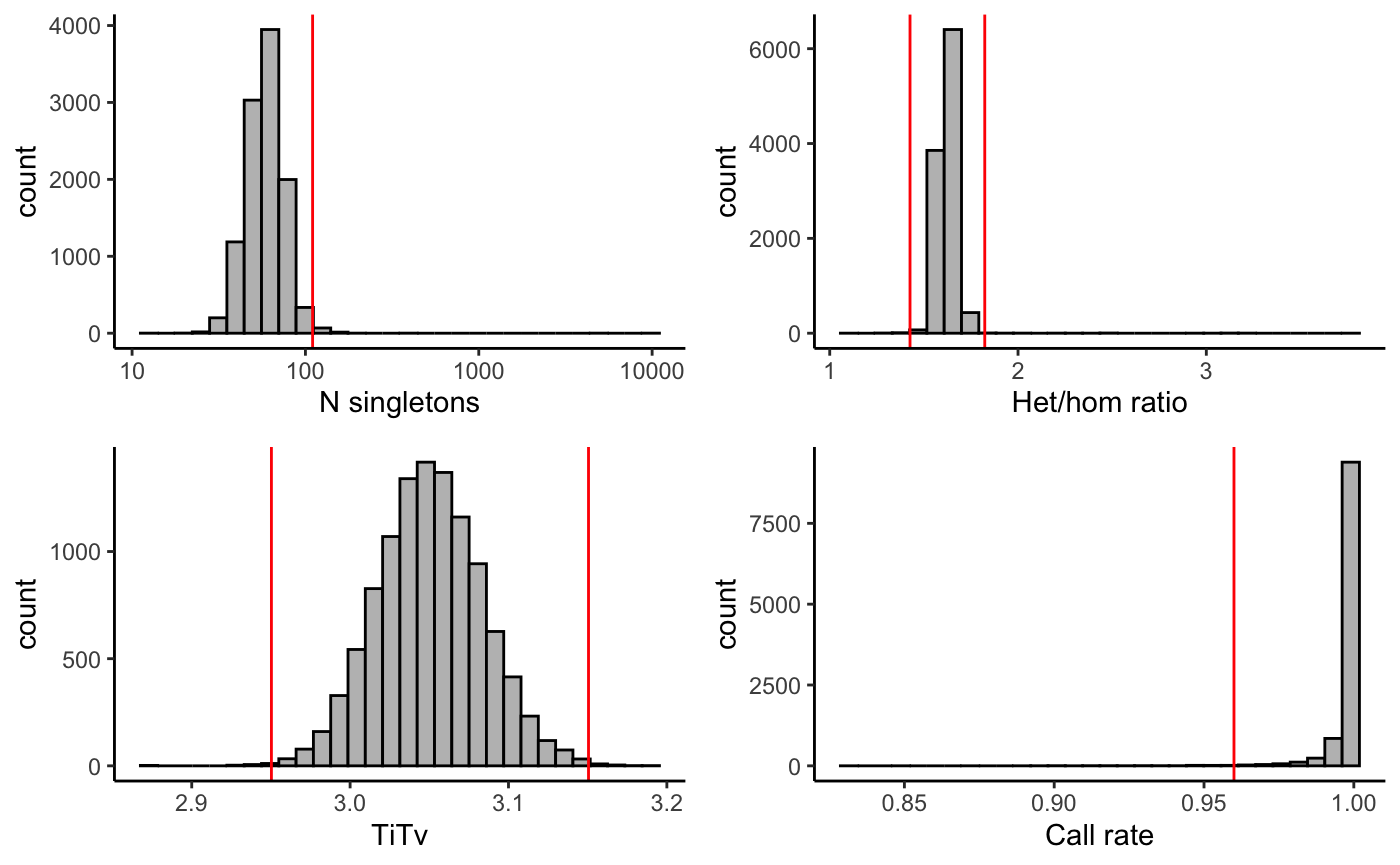

**Supplementary Figure 3.** Distributions of sample-level QC metrics in new case-control sample. Histograms show distribution of the number of singleton variants, heterozygous/homozygous variant ratio, transition/transversion ratio, and call rate per sample. Red lines indicate the threshold used to filter samples that failed ‘hard filter’ QC.

**Exclusion of samples overlapping with SCHEMA study**: To ensure that our new sample is independent from cases we contributed to the SCHEMA study, we performed an IBD analysis using microarray data from all schizophrenia datasets held by Cardiff University (CLOZUK 1, 2 and 3, CardiffCOGs, F-series and affected-sibpair samples), and excluded 6 cases in our new exome-sequencing sample found to be a duplicate (PI-HAT value > 0.9) with a case included in the SCHEMA study. Additionally, for each new case/control sample with a rare variant in the 12 previously implicated exome-wide significant schizophrenia genes, or in any of the novel risk genes identified in the current study, we examined the percentage of singleton coding variants carried across the exome that are also observed in SCHEMA cases. The mean percentage of singleton variants carried by these samples that are also observed in SCHEMA was 22.7%, with two cases being clear outliers with > 75% of their singleton variants also observed in SCHEMA (Supplementary Figure 4). To ensure that our primary findings are not influenced by sample overlaps, we excluded these two cases from our study.

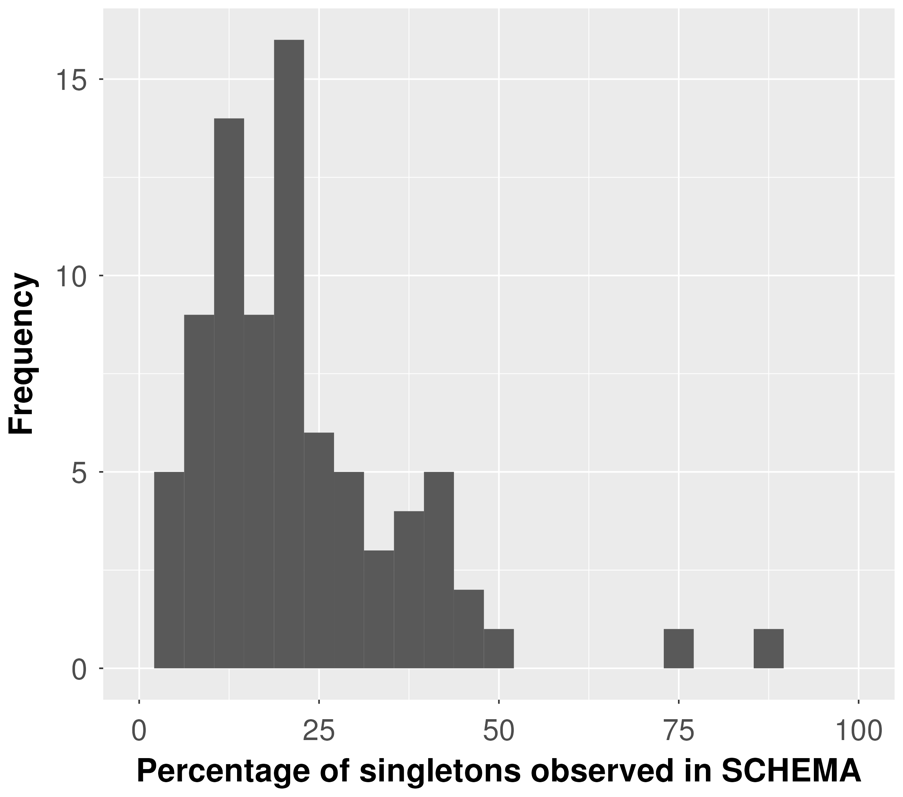

**Supplementary Figure 4.** Percentage of variants per-sample observed in SCHEMA. For each of our new samples with a rare variant in the 12 previously implicated exome-wide significant schizophrenia genes, or in any of the novel risk genes identified in the current study, the percentage of singleton coding variants carried across the exome that are also observed in SCHEMA cases is shown.

**Contamination:** For samples passing the above QC, we estimated sequencing contamination using the compute_charr method in Hail^14^. All samples had < 5% estimated contamination. Thus, no samples were excluded for having excess contamination.

**Summary of all sample quality control**

After applying all sample-level quality control described above, 4,650 cases and 5,719 controls were retained for analysis. A hierarchical summary of sample exclusions is provided in Supplementary Table 1.

|  | **Control** | **Schizophrenia** |
| --- | --- | --- |
| Count pre-QC | 7,268 | 5,525 |
| Initial sample exclusions | 4 | 24 |
| Sex check exclusions | 133 | 37 |
| Relatedness exclusions | 331 | 224 |
| Ancestry exclusions | 1,001 | 418 |
| Hard filter exclusions | 80 | 164 |
| SCHEMA sample overlaps | 0 | 8 |
| **Passed samples** | **5,719** | **4,650** |
| **Passed males** | **2,729** | **3,329** |
| **Passed females** | **2,990** | **1,321** |

**Supplementary Table 1.** Hierarchical summary of quality control sample exclusions in the new case-control sample.

#### Genotype-level quality control

Genotype calls were defined as low quality and excluded if they met any of the following criteria: total depth < 10X; genotype quality score < 20; allele balance > 0.1 and < 0.9 for homozygous genotypes for the reference and alternative allele, respectively; allele balance < 0.25 or > 0.75 for heterozygous genotypes. For variants falling within the X chromosome non-pseudoautosomal (non-PAR) region, male heterozygote genotypes were excluded.

#### Variant-level quality control

Variant sites were defined as low quality and excluded if they met any of the following criteria: number of observed alleles > 6; in a region of low sequence complexity^15^; failed GATK VQSR filters; not in Hardy-Weinberg equilibrium (P-value < 10^−8^). To account for differences in sequencing depth between cases and controls, variant sites were excluded if they had call rate < 0.97 in the full sample or in the cases, the general controls, or in the AD samples only.

#### Variant annotation

Variants were annotated with their most severe consequence across transcripts using the Ensembl Variant Effect Predictor (version 96; McLaren et al. 2016). PTVs were defined as nonsense, splice-site or frameshift variants. Missense variants were annotated using the missense badness, Polyphen-2 and constraint (MPC) score^17^.

#### Analysis of SCHEMA rare coding variant data

We re-analysed RCVs from 24,248 cases and 97,322 controls in the SCHEMA study using variant level results obtained from the SCHEMA Browser (<https://schema.broadinstitute.org/downloads>). SCHEMA case-control variants are derived from 12 sample collections and 46,885 external gnomAD controls, which the SCHEMA study stratified into 11 independent groups based on ancestry and exome capture platform (Supplementary Table 2). We analysed RCVs in the SCHEMA cases and controls using Cochran-Mantel-Haenszel (CMH) tests with continuity correction, with separate contingency tables for the 11 SCHEMA strata. Per-gene CMH P-values for synonymous variants (MAC ≤ 5, which is the same MAC used in the SCHEMA study) followed the expected null distribution (Supplementary Figure 5), suggesting our CMH tests are well-controlled for stratification within the SCHEMA case-control sample.

| **Stratum** | **SCHEMA Cases** | **SCHEMA controls** | **gnomAD controls** |
| --- | --- | --- | --- |
| EUR (Exomes, Nextera) | 8874 | 19074 | 23561 |
| EUR (Exomes, non-Nextera) | 7277 | 11187 | 0 |
| AMR (Exomes, Nextera) | 1388 | 3146 | 12008 |
| FIN (Exomes, non-Nextera) | 944 | 7984 | 3542 |
| EAS (Exomes, non-Nextera) | 1730 | 1607 | 6806 |
| AFR (Whole Genomes) | 2245 | 1170 | 420 |
| ASJ (Exomes, Nextera) | 869 | 2415 | 548 |
| EST (Whole Genomes) | 261 | 2281 | 0 |
| FIN (Whole Genomes) | 423 | 655 | 0 |
| AFR (Exomes, non-Nextera) | 127 | 765 | 0 |
| SAS (Exomes, non-Nextera) | 110 | 153 | 0 |

**Supplementary Table 2**: SCHEMA sample counts across 11 strata defined by ancestry and exome capture platform. Strata description and sample numbers were taken from Singh et all 2022^18^.

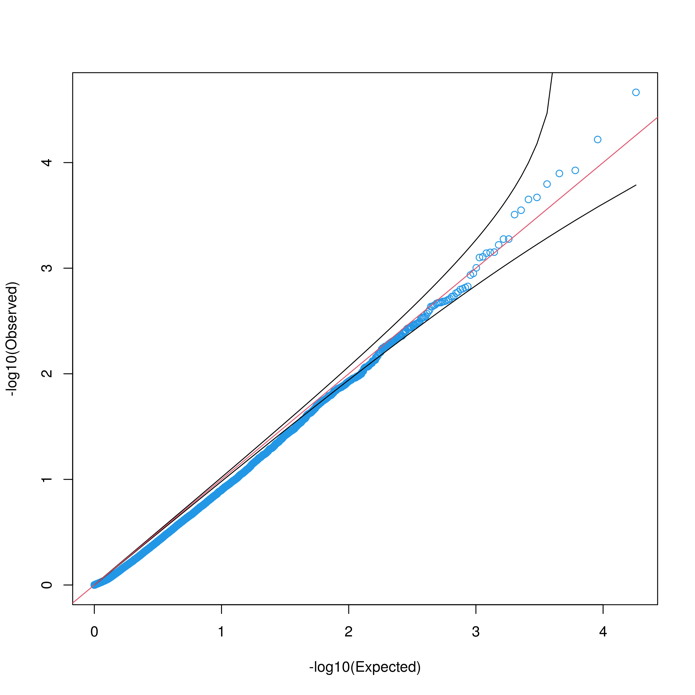

**Supplementary Figure 5**. QQ plot showing per-gene synonymous variant P-values generated using Cochran-Mantel-Haenszel tests over the 11 SCHEMA case-control strata.

#### Gene set analysis in the new case-control sample

Gene set enrichment analysis was performed in the new case-control sample using Firth’s Penalised logistic regression (logistf function in R), where case-control status was regressed on the number of rare coding variants in a given gene set, controlling for the first ten PCs derived from common variants in the exome-sequencing data, sex, and the exome-wide burden of singleton synonymous variants.

We compared effect sizes between independent gene-set enrichment tests using the following z-test equation:

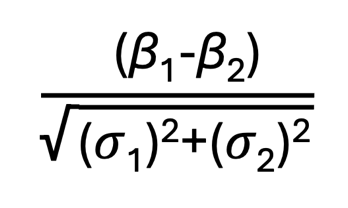

Where β_1_ represents the beta and σ_1_ the standard error for burden of variants for gene-set 1, and β_2_ represents the beta and σ_2_ the standard error for burden of variants in gene-set 2.

#### Replication of single genes in the new sample

In our new case-control sample, we performed a single-gene analysis for 12 genes previously associated with rare coding variants in schizophrenia at exome-wide significance. We restricted this analysis to variants with a MAC ≤ 5 in the new case-control sample and a MAC ≤ 5 in the control subsample of gnomAD^19^, which closely aligns with the MAC used in the SCHEMA study. For each gene, we tested the class of variant most strongly associated with schizophrenia in our re-analysis of SCHEMA case-control RCVs (described above). Since this replication analysis only involved our new sample, which is homogenous in terms of genetically inferred ancestry and sequencing platform, association statistics were generated for autosomal and pseudoautosomal genes using one-sided Fisher’s exact tests. For non-pseudoautosomal genes on the X chromosome, association statistics were generated using one-sided CMH tests, with separate contingency tables for males and females.

### 2. Relationship between sequencing depth and burden of singleton coding variants

In our new sample, sequencing coverage was greater in controls (mean genotype coverage = 34.2x) than in cases (mean genotype coverage = 26.0x). To evaluate whether this coverage difference impacts our case-control RCV enrichment analysis, we examined the relationship between sequencing depth and the rate of singleton (minor allele count = 1 and absent in gnomAD controls^20^) coding variants in our new sample. We did this using linear regression models, where the number of singleton coding variants in LoF-tolerant genes is the dependent variable, and the per-sample mean genotype depth is the independent variable, controlling for 10 PCs and sex. We found that the collective burden of synonymous variants, missense variants and PTVs was positively correlated with mean genotype depth (Supplementary Table 3). Moreover, when tested independently, both synonymous variants and missense variants were associated with mean genotype depth (Supplementary Table 3). These findings suggest that the control excess of synonymous variants in LoF intolerant genes that we observed in our new sample (Supplementary Table 4) is likely driven by the greater sequencing coverage in controls compared with cases. Thus, higher control sequencing coverage may produce more conservative estimates of schizophrenia rare variant enrichment in our new sample.

| **Variant class** | **Mean genotype depth** | |
| --- | --- | --- |
|  | **Beta** | **P value** |
| Synonymous variants + missense variants + PTVs | 0.026 | 0.0034 |
| Synonymous | 0.011 | 0.0068 |
| Missense | 0.014 | 0.044 |
| PTVs | 0.0016 | 0.49 |

**Supplementary Table 3**. Relationship between sequencing depth and burden of singleton rare coding variants in the new case-control sample. Beta coefficients and P values are derived from linear regression models, where the number of singleton coding variants in LoF-tolerant genes is the dependent variable, and the per-sample mean genotype depth is the independent variable, controlling for 10 PCs and sex. P values are two sided.

### 3. Summary of novel schizophrenia FDR < 5% genes.

###### SLC6A1

*SLC6A1* encodes the gamma-aminobutyric acid (GABA) transporter GAT1, which is expressed in neurons and mediates uptake of GABA from the synaptic cleft of inhibitory synapses. As a protein marker for GABAergic neurons, GAT1 shows the strongest immunoreactivity for parvalbumin-expressing cortical interneurons^21^, which synapse on the axon initial segments of pyramidal neurons and play a key role in synchronising glutamatergic outputs at the gamma frequency to facilitate cognitive processes such as information processing and working memory^22,23^. In schizophrenia, these inhibitory neurons show consistent hypoactivity and loss of gamma-oscillatory activity^24,25^, which is accompanied by decreased expression of GAT1^26,27^, in addition to other features such as reduced activity of glutamic acid decarboxylase (GAD) and lower concentrations of GABA^28^. Recently published *in vitro* GABA uptake assay data suggests schizophrenia *SLC6A1* missense variants confer loss-of-function effects on GAT-1 protein leading to reduced GABA uptake^29^. A previous schizophrenia CNV study also found genes related to the GABA_A_ receptor complex gene set are enriched for both deletions and duplications in cases compared to controls^30^.

Missense variants and PTVs in *SLC6A1* also confer risk to other psychiatric and developmental disorders. A set of *SLC6A1*-related disorder (SRD) patients has been identified from clinical cohorts of ID and childhood epilepsy; SRD is typically characterised by myoclonic-atonic epilepsy, mild or moderate ID, and autistic features^31–33^. *SLC6A1* is also significantly implicated in rare variant association studies of DD/ID and autism^34,35^.

###### PCLO

*PCLO* encodes for Piccolo, a component of the presynaptic cytoskeletal matrix at glutamatergic and GABAergic synapses^36^. Piccolo is thought to act as a scaffold protein promoting formation of the synapse and coordination of synaptic vesicles at the active zone where neurotransmitters are released^37^. Complete loss-of-function of *PCLO* leads to pontocerebellar hypoplasia type III in humans^38^ and in rat models^39^, which in humans encompasses global developmental delay and seizures. In the PGC3SEQ targeted sequencing meta-analysis, *PCLO* was implicated as a shared risk gene between schizophrenia and ASD^40^.

###### ZMYND11

*ZMYND11* encodes a chromatin remodelling protein which recognises H3.3K36me3 marks at actively transcribed genes and acts to inhibit transcriptional elongation^41^. The H3.3 histone variant has been shown to accumulate in the neuronal genome and to be particularly relevant for expression of synaptic genes^42^. *ZMYND11* has also been shown to contribute to regulation of neuronal differentiation^43^. Mutations in *ZMYND11* are associated with syndromic intellectual disability, encompassing developmental delay, epilepsy, and features of ASD and ADHD^44,45^. The majority of identified mutations are protein-truncating, with a small number of missense and other mutations; missense variants have also been identified in two patients with unusually severe phenotypes, which were suspected to lead to gain-of-function effects^46,47^.

###### BSCL2

*BSCL2* encodes seipin, an endoplasmic reticulum (ER)-localised protein which mediates formation of lipid droplets as an energy store within the cell ^48^. PTVs in *BSCL2* are associated with severe lipodystrophy encompassing diabetes and intellectual disability, and gain-of-function variants are associated with a spectrum of neuropathic conditions including distal hereditary motor neuropathy, Silver syndrome, and spastic paraplegia^49^. Splice variants in *BSCL2* have also been suggested to cause epileptic encephalopathy^50^ and ASD with parkinsonism^51^.

###### KLC1

*KLC1* codes for a light chain subunit of kinesin, a tetrameric protein complex responsible for intracellular transport along the cytoskeleton. Two heavy chains function as motors, while two light chains act as adaptors binding cargo to be transported^52^. In neurons, *KLC1* has been shown to play a role in vesicle transport through its interaction with Calsyntenin-2^53^, and knockdown of *KLC1* in a human cell line was found to impair neuronal differentiation^54^. In addition to implication of *KLC1* in schizophrenia by the largest GWAS to date^55^, increased expression of *KLC1* was found to be significantly associated with risk in a schizophrenia TWAS^56^.

###### CGREF1

Lastly, *CGREF1* encodes an extracellular protein thought to play a role in cell proliferation^57^, and has previously been identified as an ASD risk gene^35^.

### 4. Supplementary Tables 4-7

| **Gene set** | **Variant class** | **N variants (rate)** | | **OR (95% CI)** | **P** |
| --- | --- | --- | --- | --- | --- |
|  |  | **Cases** | **Controls** |  |  |
| Constrained genes  (n = 3,051) | PTV | 869 (0.19) | 970 (0.17) | 1.14 (1.03-1.27) | 1.1 x 10^-2^ |
|  | MPC > 3 | 70 (0.015) | 61 (0.011) | 1.59 (1.08-2.33) | 1.8 x 10^-2^ |
|  | MPC 2-3 | 1068 (0.23) | 1346 (0.24) | 1.04 (0.94-1.14) | 0.45 |
|  | Synonymous | 6764 (1.5) | 9990 (1.7) | 0.93 (0.89-0.96) | 2.5 x 10^-5^ |

Supplementary Table 4: Gene-set analysis of singleton coding variants in constrained genes in the new case-control sample. Odds ratios (OR) were derived from Firth’s logistic regression models (Supplementary Methods). Constrained genes are defined as those with pLi scores ≥ 0.9 in gnomAD^19^. PTV = protein-truncating variants; MPC = ‘missense badness, Polyphen-2 and constraint’ score.

| **Gene set** | **Variant class** | **N variants (rate)** | | **OR (95% CI)** | **P** |
| --- | --- | --- | --- | --- | --- |
|  |  | **Cases** | **Controls** |  |  |
| Published exome-wide significant genes (n = 12) | PTV | 25 (0.0054) | 7 (0.0012) | 4.93 (2.04-13.16) | 2.8 x 10^-4^ |
|  | MPC >3 | 5 (0.0011) | 7 (0.0012) | 0.86 (0.22-3.02) | 0.82 |
|  | MPC 2-3 | 18 (0.0039) | 21 (0.0037) | 1.20 (0.57-2.51) | 0.63 |
|  | Synonymous | 223 (0.048) | 295 (0.052) | 0.94 (0.76-1.15) | 0.53 |
| Published FDR < 5% genes  (n = 20) | PTV | 21 (0.0045) | 16 (0.0028) | 2.46 (1.18-5.15) | 1.6 x 10^-2^ |
|  | MPC >3 | 0 (0) | 0 (0) | NA | NA |
|  | MPC 2-3 | 21 (0.0045) | 28 (0.0049) | 0.98 (0.51-1.86) | 0.95 |
|  | Synonymous | 230 (0.049) | 293 (0.051) | 0.96 (0.79-1.18) | 0.72 |

Supplementary Table 5. Gene-set analysis of previously implicated genes in the new case-control sample. Variants are restricted to those with a minor allele count ≤ 5 in the new sample and ≤ 5 in gnomAD controls. Odds ratios (OR) were derived from Firth’s logistic regression models (Supplementary Methods). Published exome-wide significant genes were derived from^18,40^. Additional published FDR < 5% genes were derived from^18^. PTV = protein-truncating variants; MPC = ‘missense badness, Polyphen-2 and constraint’ score.

| Gene set | Gene-set enrichment | | | Z-test P value |
| --- | --- | --- | --- | --- |
|  | **beta** | **SE** | **P** |  |
| 12 previously implicated exome-wide significant genes | 1.59 | 0.467 | 2.8 x 10^-4^ | 7.6 x 10^-4^ |
| Independent set of constrained genes | 0.108 | 0.0445 | 0.016 |  |
| 20 additional previously implicated genes at FDR < 5% | 0.898 | 0.372 | 0.016 | 0.017 |
| Independent set of constrained genes | 0.108 | 0.0445 | 0.016 |  |

Supplementary Table 6. Comparison of effect sizes for PTVs in previously implicated genes and constrained genes. Variants are restricted to PTVs with a minor allele count ≤ 5 in the new sample and ≤ 5 in gnomAD controls. Beta, standard errors (SE) and P-values are derived from a gene-set analysis in our new case-control sample. See Supplementary Methods for a description of the Z-test used to compare gene-set enrichment statistics.

| **Gene symbol** | **Variant class** | **New sample** | | | |
| --- | --- | --- | --- | --- | --- |
|  |  | **Case variants** | **Control variants** | **OR (95% CI)** | **P-val** |
| ***SETD1A*** | PTV + MPC>2 | 8 | 2 | 4.9 (1.0-47.6) | 0.027 |
| ***XPO7*** | PTV + MPC>3 | 4 | 0 | Inf (0.8-Inf) | 0.040 |
| ***HERC1*** | PTV | 4 | 1 | 4.9 (0.5-242.2) | 0.13 |
| ***TRIO*** | PTV | 0 | 1 | 0.0 (0.0-47.9) | 1.00 |
| ***SP4*** | PTV + MPC>3 | 7 | 5 | 1.7 (0.5-6.9) | 0.26 |
| ***GRIN2A*** | PTV + MPC>2 | 3 | 2 | 1.8 (0.2-22.1) | 0.40 |
| ***CACNA1G*** | PTV + MPC>3 | 0 | 4 | 0.0 (0.0-1.9) | 1.00 |
| ***CUL1*** | PTV + MPC>3 | 0 | 1 | 0.0 (0.0-47.9) | 1.00 |
| ***AKAP11*** | PTV | 3 | 0 | Inf (0.5-Inf) | 0.090 |
| ***SRRM2*** | PTV | 5 | 0 | Inf (1.1-Inf) | 0.018 |
| ***GRIA3*** | PTV + MPC>3 | 0 | 0 | 0.0 (0.0-Inf) | 1.00 |
| ***RB1CC1*** | PTV | 2 | 1 | 2.5 (0.1-145.1) | 0.42 |

Supplementary Table 7. Analysis of previously reported exome-wide significant genes in the new case-control data. Variant counts, odds ratios (ORs) and P-values correspond to the variant class shown. Variant classes and ordering correspond to SCHEMA re-analysis P-values (Extended Data Table 2). P-values are 1-sided. ORs are based on 2-sided tests to produce upper-bound confidence interval (CI) estimates.

### 5. Gene discovery sensitivity analysis

A sensitivity analysis was performed to determine whether our novel genes remained significant after excluding individuals with Alzheimer’s disease from our new control sample. In this sensitivity analysis, both *STAG1* and *ZNF136* remained associated at exome-wide significance, and among the 6 additional FDR < 5% significant novel genes, only *CGREF1* is no longer significant at FDR < 5% (*CGREF1*: Q-value = 0.056; Supplementary Table 8).

| **Gene symbol** | **Variant class** | **Case-control-de novo variant meta-analysis** | |
| --- | --- | --- | --- |
|  |  | **P-val** | **Q-val** |
| *STAG1* | PTV + MPC >2 | **1.5 x 10^-7^** | 4.6E-04 |
| *ZNF136* | PTV | **1.5 x 10^6^** | 0.0031 |
| *SLC6A1* | MPC >2 | 3.6 x 10^6^ | 0.0046 |
| *KLC1* | MPC >2 | 1.1 x 10^5^ | 0.012 |
| *PCLO* | PTV | 2.4 x 10^5^ | 0.024 |
| *ZMYND11* | PTV | 2.5 x 10^5^ | 0.024 |
| *BSCL2* | PTV | 5.3 x 10^5^ | 0.043 |
| *CGREF1* | PTV | 8.3 x 10^5^ | 0.061 |

Supplementary Table 8. Sensitivity analysis of exome-wide significant and FDR < 5% genes. Case-control-*de novo* variant enrichment statistics are presented after excluding individuals with Alzheimer’s disease from the new control sample. Bold text indicates P-values exceeding Bonferroni significance (P < 1.63 x 10^-6^). P-values are two sided. Q-values show adjusted P-values using the false discovery rate approach. PTV = protein-truncating variant; MPC = ‘missense badness, Polyphen-2 and constraint’ scores for missense variants.

### 6. Extended Data Table Legends

**Extended Data Table 1**: Variants in new case-control sample that contribute to novel exome-wide significant and FDR < 5% genes. Locus coordinates are in build37. MPC = ‘missense badness, Polyphen-2 and constraint’ score.

**Extended Data Table 2**: Full gene case-control and *de novo* enrichment results. SCHEMA case-control variant counts are derived from our reanalysis of variant level data downloaded from the SCHEMA Browser (<https://schema.broadinstitute.org/downloads>. See Methods for a full description of how the combined case-control statistics were generated. In the combined case-control statistics columns, dashes correspond to tests not passing the two-strata adjustment, and NAs correspond to genes which do have any potential missense MPC>2 or MPC>3 variants (see Methods). *De novo* variant counts were derived from 3,444 published schizophrenia trios^58^. In the ‘*de novo* variant P values’ columns, results are only shown for tests which had a combined case-control p-value < 0.01 (see Methods for further detail). The ‘Final case-control-de novo meta-analysis’ columns show the most significant result for all tests performed for each gene, and the class of mutation corresponding to the most significant result. Q-values are derived from the distribution of P values across all single gene tests performed (see Methods), and not just the P value distribution given in the “Final case-control-de novo meta-analysis” column.
