## Extended data figure 1 for "Analysis of exome sequencing data implicates rare coding variants in *STAG1* and *ZNF136* in schizophrenia"

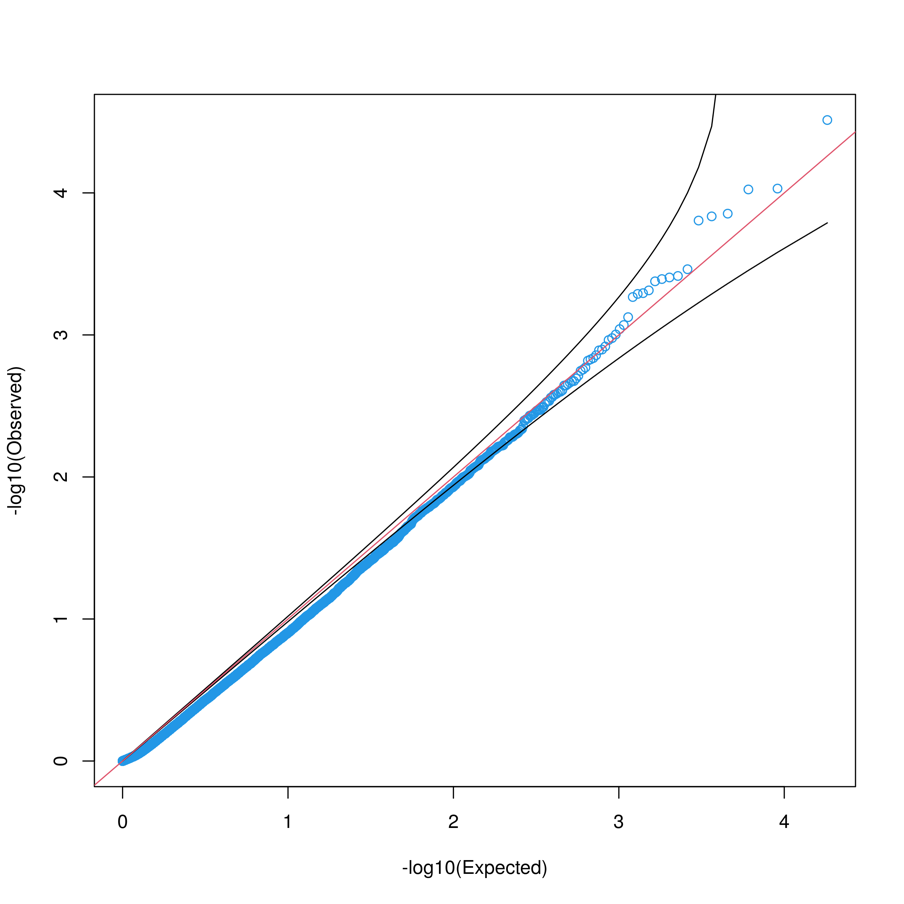

**Extended Data Figure 1.** QQ plot for synonymous variant P-values in the combined case-control analysis (SCHEMA + new sample). P-values are 2-sided and were generated using Cochran-Mantel-Haenszel tests with 12 contingency tables for autosomal and pseudoautosomal genes and 13 contingency tables for non-pseudoautosomal genes (Full approach described in Methods).
